# Duration of Supplemental Oxygen Requirement and Predictors in Severe COVID-19 Patients in Ethiopia: A Survival Analysis

**DOI:** 10.1101/2020.10.08.20209122

**Authors:** Tigist W. Leulseged, Ishmael S. Hassen, Mesay G. Edo, Daniel S. Abebe, Endalkachew H. Maru, Wuletaw C. Zewde, Nigat W. Chamiso, Tariku B. Jagema

## Abstract

**Background:** With the rising number of new cases of COVID-19, understanding the oxygen requirement of severe patients assists in identifying at risk groups and in making an informed decision on building hospitals capacity in terms of oxygen facility arrangement. Therefore, the study aimed to estimate time to getting off supplemental oxygen therapy and identify predictors among COVID-19 patients admitted to Millennium COVID-19 Care Center in Ethiopia.

**Methods:** A prospective observational study was conducted among 244 consecutively admitted COVID-19 patients from July to September, 2020. Kaplan Meier plots, median survival times and Log-rank test were used to describe the data and compare survival distribution between groups. Cox proportional hazard survival model was used to identify determinants of time to getting off supplemental oxygen therapy, where hazard ratio (HR), P-value and 95%CI for HR were used for testing significance and interpretation of results.

**Results:** Median time to getting off supplemental oxygen therapy among the studied population was 6 days (IQR, 4.3-20.0). Factors that affect time to getting off supplemental oxygen therapy were age group (AHR=0.52,95%CI=0.32,0.84, p-value=0.008 for ≥70 years) and shortness of breath (AHR=0.71,95%CI=0.52,0.96, p-value=0.026).

**Conclusions:** Average duration of supplemental oxygen therapy requirement among COVID-19 patients was 6 days and being 70 years and older and having shortness of breath were found to be associated with prolonged duration of supplemental oxygen therapy requirement. This result can be used as a guide in planning institutional resource allocation and patient management to provide a well-equipped care to prevent complications and death from the disease.

## INTRODCUTION

The 2019 coronavirus disease was first identified in China on December 2019 and later spread to the entire world and declared to be a pandemic by world health organization on March 11, 2020 (1). In Ethiopia the first case was identified on March 13, 2020, two days after the pandemic was declared. According to the national report, as of January 25, 2020, a total of 134,132 cases were identified with 120,199 recovered cases 2071 deaths. Since the disease transmission dynamic shifted to a community transmission, the number of new cases, those who need critical care and daily deaths are increasing, with 231 cases on critical care at different treatment centers in the country on the same day (2). Considering the countries overburdened health care system, providing adequate care for COVID-19 patients who need hospitalization and especially supplemental oxygen therapy might be challenging if the pandemic continues.

To halt the transmission of the dynamics and because of limited knowledge on the disease progression and outcome, at first every patient who tested positive for SARS-COV-2 used to be quarantined and observed till Virological recovery. But eventually as the number of case increases the admission and discharge criteria changed in order to accommodate the service to those who needs it most. and This criteria was also applied in our country (3, 4).

Assessing the resources needed for admitted patients can be guided by the length of hospital stay or time needed to get virological recovery or clinical improvement. So far studies that assessed improvement from the disease in terms of length of hospital stay (5-12) and viral shedding duration (13-17) were conducted and showed a wide difference in reported values ranging from few days up to a couple of months or more. Assessing the disease recovery in terms of clinical parameters provides better detail about the disease effect on the patient and its burden on the health care system compared to assessing the length of stay or virological recovery per se as it also tells us the associated resource requirement during the hospital stay. In other words, a patient staying at a ward with no need of oxygen therapy or expensive antibiotics and someone on intensive care doesn’t cost the same to the institution. This implies that, hospital length of stay or duration of viral shedding might not directly correlate with disease severity and outcome. Thereby, using these two parameters have minimal cost implication to the health care system in the current practice compared to using clinical criteria like having symptomatic disease or need of medication or supplemental oxygen therapy.

Measuring duration of supplemental oxygen requirement helps not only in identifying who is at most risk of prolonged oxygen support need and should be protected but it also assists in deciding on building hospitals capacity in terms of oxygen facility arrangement and organizing ICU and wards with inbuilt supplemental oxygen system. Therefore, in this study we aimed to assess severe COVID-19 patients’ clinical improvement by measuring the time needed from admission and being on supplemental oxygen therapy to getting off supplemental oxygen therapy and its predictors among patients admitted to Millennium COVID-19 Care Center in Ethiopia.

## MATERIALS AND METHODS

### Study setting

The study was conducted at Millennium COVID-19 Care Center (MCCC), a makeshift hospital in Addis Ababa, the capital city of Ethiopia. MCCC is remodeled from the previous Millennium hall which was a recreational center. The Center started accepting patients on June 2, 2020 with a Capacity of 1000 Beds, 40 ICU beds and 12 mechanical ventilators. Currently the clinical team is composed of 10 emergency physicians, 8 internists, 2 surgeons, 33 General Practitioners, 176 nurses, 8 pharmacists, 15 laboratory technologists, 1 radiologist and 6 radiology technologists.

### Study Design and Population

The study design was hospital based prospective observational design. The observation was made from July to September, 2020.

The source population was all severe cases of COVID-19 admitted at MCCC with a confirmed diagnosis of COVID-19 using RT-PCR, as reported by a laboratory given mandate to test such patients by the Ministry of Health and who were on follow up from July to September, 2020 (3).

During this interval a total of 244 Severe COVID-19 patients were admitted to the Center. All the 244 patients were included in the study.

### Sample size Determination and Sampling Technique

All consecutively admitted Severe COVID-19 patients during the follow up period were included in the study.

### Eligibility criteria

All Severe COVID-19 patients who were on treatment and follow up at the MCCC from July to September, 2020 were included.

### Operational Definitions

#### COVID-19 severity

was determined based on the WHO classification as follows (4).

- **Mild Disease:** Characterized by fever, malaise, cough, upper respiratory symptoms, and/or less common features of COVID-19 (headache, loss of taste or smell etc.)
- **Moderate Disease:** Patients with lower respiratory symptom/s. They may have infiltrates on chest X-ray. These patients are able to maintain oxygenation on room air.
- **Severe Disease:** These patients have developed complications. The following features can define severe illness.
  - Hypoxia: SPO2 ≤ 93% on atmospheric air or PaO2:FiO2 < 300mmHg (SF ratio < 315)
  - Tachypnea: in respiratory distress or RR>30 breaths/minutes
  - More than 50% involvement seen on chest imaging

#### Event

Getting off supplemental oxygen therapy as decided by maintaining oxygen saturation level of above 93% on atmospheric air measured both at rest and while ambulating.

#### Censoring

Includes patients lost to follow-up, transferred out, died or completed the follow-up period before getting off supplemental oxygen therapy.

Time to event or censoring: time between initiations of supplemental oxygen therapy to getting off oxygen supplement (in days).

### Data Collection Procedures and Quality Assurance

An interviewer administered pretested questionnaire that consists of the variables of interest was developed from the patient registration and follow up form and used to collect the necessary data from the patients and their medical charts.

Training on the basics of the questionnaire and data collection tool was given for ten data collectors (Bsc nurses and General practitioners) and two supervisors (General practitioner and public health specialist) for one day.

Data consistency and completeness was checked before an attempt was made to enter the code and analyze the data.

#### Data Management and Data Analysis

The collected data was coded and entered into Epi-Info version 7.2.1.0, cleaned and stored and exported into SPSS version 23 for analysis. Frequency tables, Kaplan Meier (KM) plots and median survival times were used to describe the data. Survival experience of different groups was compared using KM survival curves. Log-rank test was used to assess significant difference among survival distributions of groups for equality.

Univariate analysis was performed to calculate an unadjusted hazard ratio (HR) and to screen out potentially significant independent variables at 25% level of significance. Association between the most relevant independent variables and the time to getting off supplemental oxygen therapy was assesses using multivariable Cox proportional hazard survival model. Adjusted HR, P-value and 95% CI for HR were used to test significance and interpretation of results. Variables with p-value ≤ 0.05 were considered as statistically associated with time to getting off supplemental oxygen therapy in days. The basic assumptions of Cox Proportional Hazard model was tested using log minus log function.

### ETHICS

The study was conducted after obtaining ethical clearance from St. Paul’s Hospital Millennium Medical College Institutional Review Board. Written informed consent was obtained from the participants. The study had no risk/negative consequence on those who participated in the study. Medical record numbers were used for data collection and personal identifiers were not used in the research report. Access to the collected information was limited to the principal investigator and confidentiality was maintained throughout the project.

## RESULT

### Socio-demographic, Co-morbid illness and drug use history, censoring status and survival experience

The mean (± SD) age of the participants was 55.1 (16.8) years. One hundred sixty five (67.6%) of the patients were males and two hundred thirty six (96.7%) of the patients were from Addis Ababa.

One hundred thirty nine (56.9%) had a history of one or more co-morbid illness. The greater proportion of the study participants were hypertensive (33.6%), followed by diabetes mellitus (28.7%), cardiac illness (11.9%) and asthma (6.1%). Thirty three (13.5%) had a history of ACEIs and/or ARBs and/or NSAID use within 14 days of admission to the center.

In all of the variable categories, the number of event achieved is greater than the censored observation. The proportion of censored observation is relatively larger as age increases, for males, for those patients with one or more pre-existing comorbid illness history and Khat chewers. On the other, those with a history of drug use had a relatively less censored observation than those with no drug use history.

The log rank test result shows that, there was a statistically significant difference in the survival time among the patients based on age group, history of pre-existing co-morbid illness, hypertension and diabetes mellitus. Accordingly, the median duration of oxygen requirement is significantly longer for those patients ≥ 70 years (22 days) followed by 50 to 69 years (8 days) and then < 50 years (6 days) (X^2^_(4)_= 15.162, P-value= 0.004). Having a history of one or more pre-existing comorbid illness (7 Vs 8 days, X^2^_(4)_= 4.449, P-value= 0.035), hypertension (7 Vs 9 days, X^2^ _(1)_= 5.204, P-value= 0.023) and diabetes mellitus (7 Vs 8 days, X^2^_(1)_ = 6.773, P-value= 0.009) resulted in a prolonged oxygen therapy requirement compared to those with no such illness.

On the other hand, the survival time did not show statistically significant difference among the patients based on sex, history of cardiac illness, asthma, khat chewing and drug use history (all p-values >0.05). (**Table 1**)

**Table 1:**
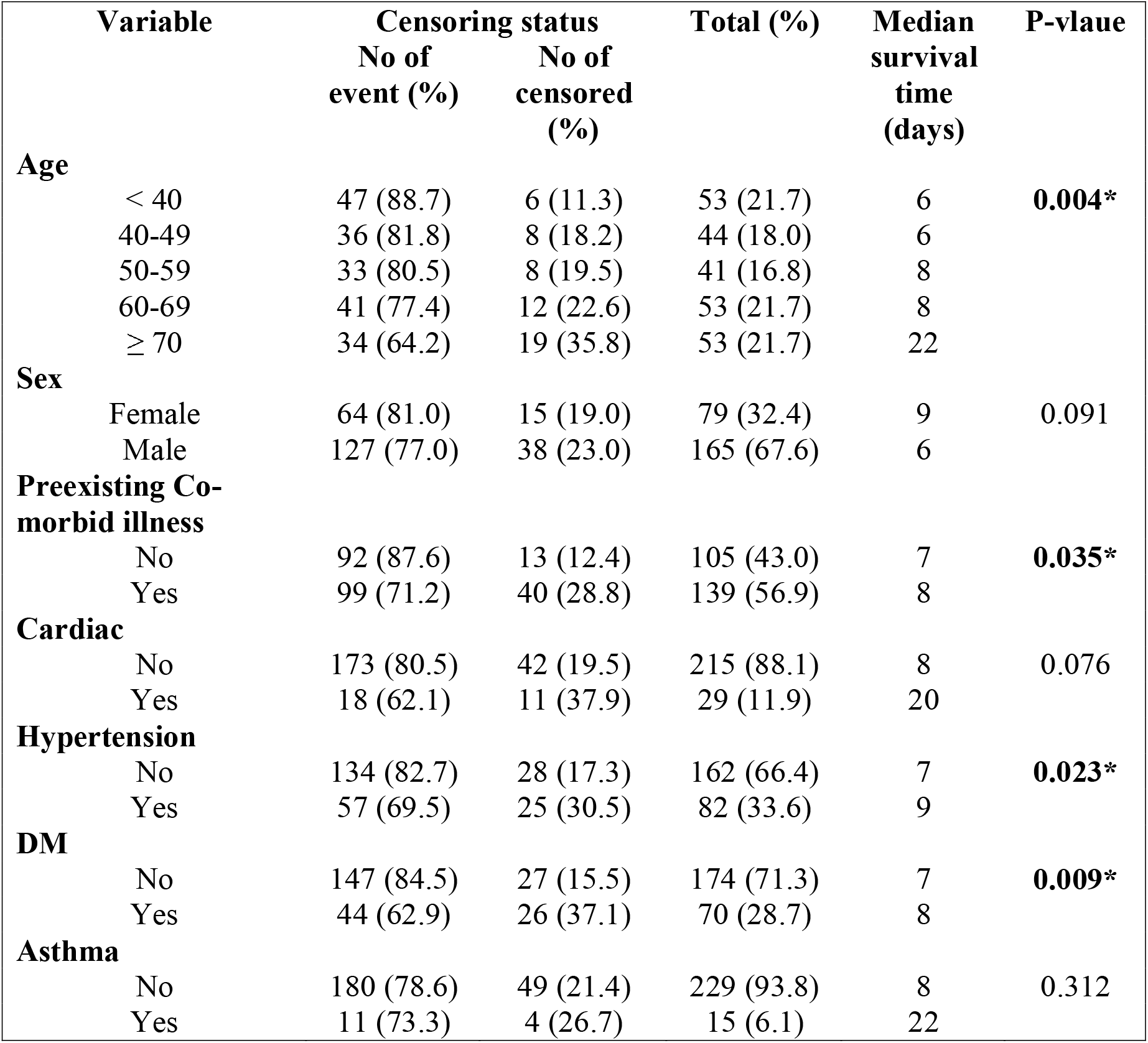
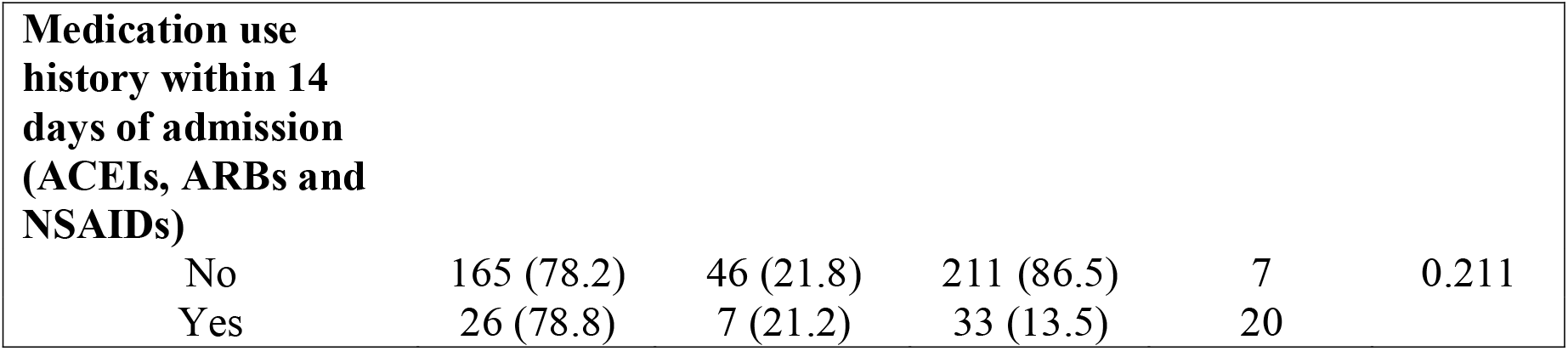
Socio–demographic, co-morbid illness and drug use history related variables censoring status and survival experience among COVID-19 patients (n=244)

| Variable | Censoring status |  | Total (%) | Median survival time (days) | P-value |
| --- | --- | --- | --- | --- | --- |
|  | No of event (%) | No of censored (%) |  |  |  |
| **Age** |  |  |  |  |  |
| < 40 | 47 (88.7) | 6 (11.3) | 53 (21.7) | 6 | **0.004\*** |
| 40-49 | 36 (81.8) | 8 (18.2) | 44 (18.0) | 6 |  |
| 50-59 | 33 (80.5) | 8 (19.5) | 41 (16.8) | 8 |  |
| 60-69 | 41 (77.4) | 12 (22.6) | 53 (21.7) | 8 |  |
| ≥ 70 | 34 (64.2) | 19 (35.8) | 53 (21.7) | 22 |  |
| **Sex** |  |  |  |  |  |
| Female | 64 (81.0) | 15 (19.0) | 79 (32.4) | 9 | 0.091 |
| Male | 127 (77.0) | 38 (23.0) | 165 (67.6) | 6 |  |
| **Preexisting Co-morbid illness** |  |  |  |  |  |
| No | 92 (87.6) | 13 (12.4) | 105 (43.0) | 7 | **0.035\*** |
| Yes | 99 (71.2) | 40 (28.8) | 139 (56.9) | 8 |  |
| **Cardiac** |  |  |  |  |  |
| No | 173 (80.5) | 42 (19.5) | 215 (88.1) | 8 | 0.076 |
| Yes | 18 (62.1) | 11 (37.9) | 29 (11.9) | 20 |  |
| **Hypertension** |  |  |  |  |  |
| No | 134 (82.7) | 28 (17.3) | 162 (66.4) | 7 | **0.023\*** |
| Yes | 57 (69.5) | 25 (30.5) | 82 (33.6) | 9 |  |
| **DM** |  |  |  |  |  |
| No | 147 (84.5) | 27 (15.5) | 174 (71.3) | 7 | **0.009\*** |
| Yes | 44 (62.9) | 26 (37.1) | 70 (28.7) | 8 |  |
| **Asthma** |  |  |  |  |  |
| No | 180 (78.6) | 49 (21.4) | 229 (93.8) | 8 | 0.312 |
| Yes | 11 (73.3) | 4 (26.7) | 15 (6.1) | 22 |  |
| **Medication use history within 14 days of admission (ACEIs, ARBs and NSAIDs)** |  |  |  |  |  |
| No | 165 (78.2) | 46 (21.8) | 211 (86.5) | 7 | 0.211 |
| Yes | 26 (78.8) | 7 (21.2) | 33 (13.5) | 20 |  |

As shown in **Figures 1** the KM survival function graph also showed that a favorable survival experience is observed among patients with no history of pre-existing co-morbid illness

**Figure 1:**
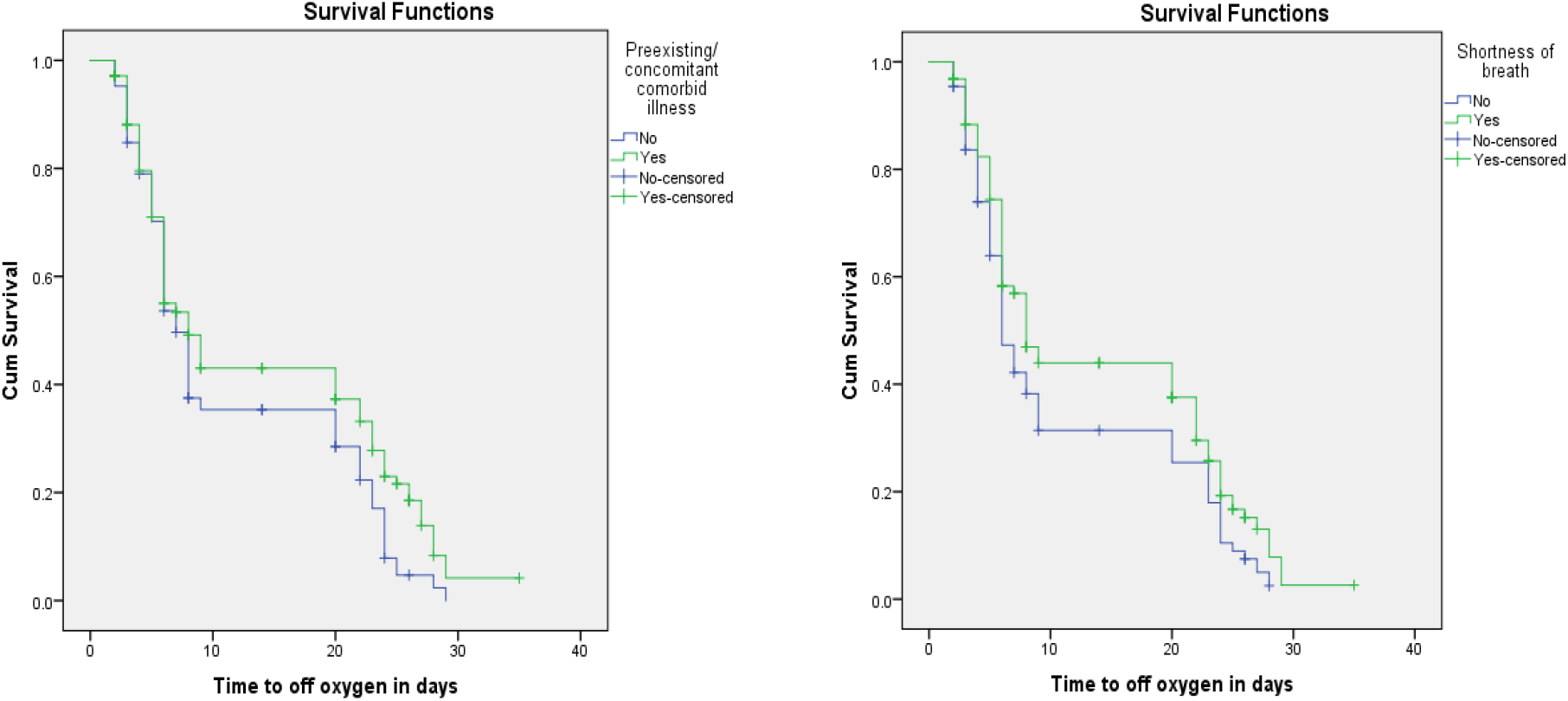
Survival and Hazard functions for pre-existing co-morbid illness and shortness of breath by time

### Presenting symptom and Oxygen saturation level related variables, censoring status and survival experience

Two hundred thirty two (95.1%) of the patients were symptomatic at presentation and the rest 12 (4.9%) of the patients had no symptom on admission other than oxygen saturation of < 93% on atmospheric air. The most frequent complaint was cough (80.7%) followed by Shortness of breath (64.3%), Cough with sputum (47.5%), Fatigue (47.5%), Fever (34.8%), Chest pain (34.1%), Headache (24.6%), Arthralgia (19.7%), Myagia (16.8%), Sore throat (16.4%), Nausea/ vomiting (7.8%) and Diarrhea (5.7%).

At admission the median oxygen saturation level on atmospheric air was 88.0% (IQR, 86-89.8).

According to the log rank test result, a statistically significant longer duration of oxygen therapy was needed among patients with a complaint of shortness of breath (8 days) compared to those with no such complaint (6 days) (X^2^_(4)_ = 4.494, P-value= 0.034). On the contrary, the log rank test didn’t show any significant difference in the survival time among the other symptom groups (all p-values >0.05). (**Table 2**)

**Table 2:**
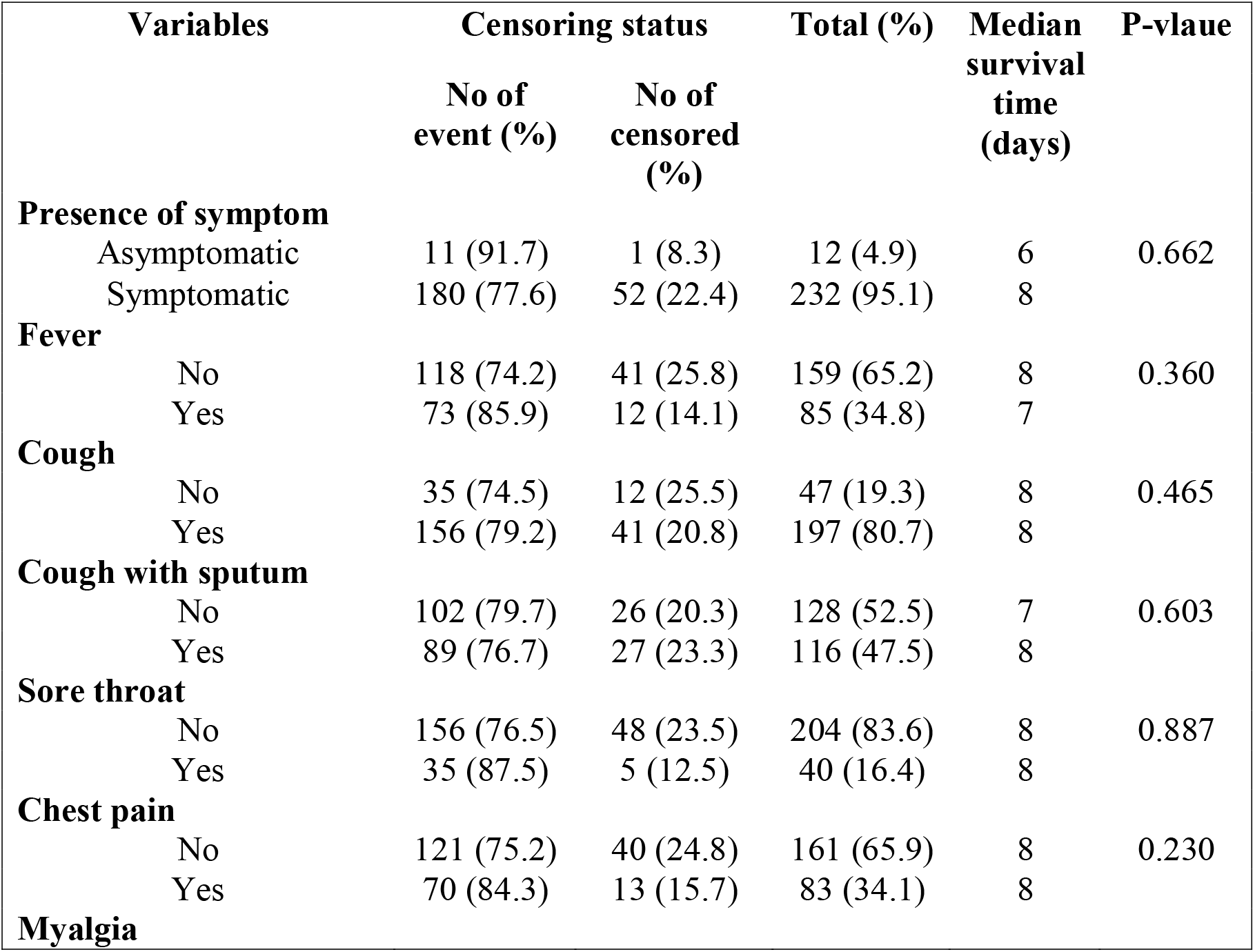
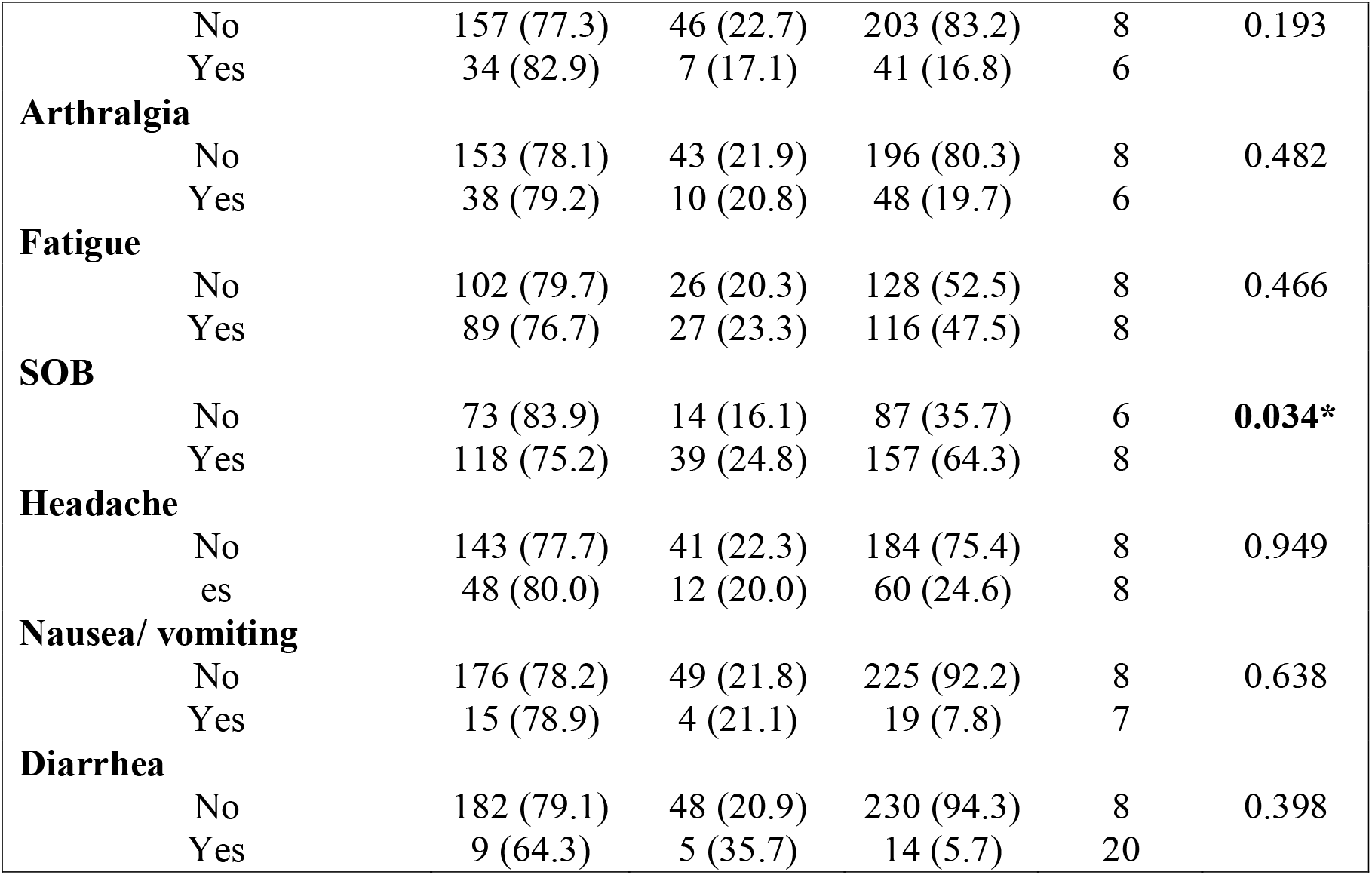
Presenting symptom related variables censoring status and survival experience among COVID-19 patients (n=244)

As shown in **Figures 1** the KM survival function graph also showed that those with no symptom of shortness of breath at admission have a favorable survival experience (time to getting off supplemental oxygen therapy) compared to those with such symptom.

### Censoring status and median time to getting off oxygen

Among the 244 patients, 191 (78.3%) of the patients achieved the event (getting off supplemental oxygen therapy) while 53 (21.7%) were censored. Among the 53 censored observation, 49 (20.1%) died and 4 (1.6%) were transferred to another hospital for further care.

The median time to getting off supplemental oxygen therapy was 6 days (IQR, of 4.3-20.0).

### Results of Multivariable Cox Proportional Hazard Model

The fundamental assumption of Cox Proportional Hazard model, which is proportional hazards assumption, was tested using Log minus Log function on SPSS version 23 software. Parallel lines between groups indicate proportionality (18). **Figures 2** shows that throughout the study time the survival curves seems to be parallel among the groups classified by age group and the presence of shortness of breath. Therefore, these plots show reasonable fit to the proportional hazard assumption.

**Figure 2:**
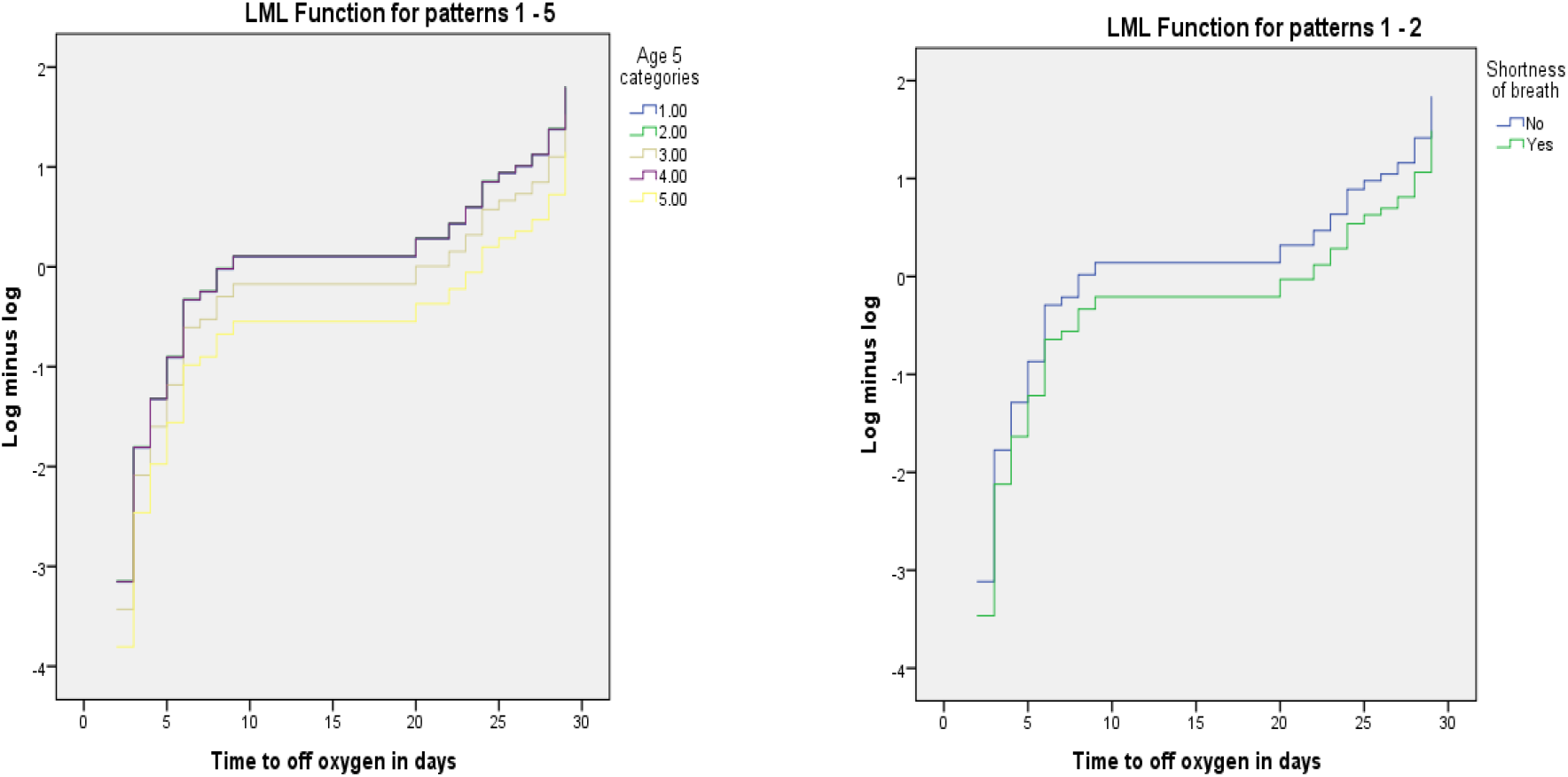
Log minus Log function for age group and shortness of breath.

Univariate analysis of each independent variable with the dependent variable was run. From univariate analysis at 25% level of significance; age group, sex, cardiac illness, hypertension, diabetes and Shortness of breath were significantly associated with duration of supplemental oxygen therapy requirement among COVID-19 patients.

However; only age group and a complaint of shortness of breath were found to be significantly associated with duration of supplemental oxygen therapy requirement in the multivariable Cox proportional hazard model at 5% level of significance.

Accordingly, after adjusting for other covariates, the rate of getting off supplemental oxygen therapy among patients ≥ 70 years was 47.8% lower than patients < 40 years old (HR= 0.52, 95% CI= 0.32, 0.84, p-value=0.008). This implies that the time needed to get off supplemental oxygen therapy was significantly longer among older patients compared with the younger patients.

Having a complaint of shortness of breath at admission was associated with a 29.5% lower rate of achieving target of off oxygen therapy compared to those patients with no such complaint on admission (HR= 0.71, 95% CI= 0.52, 0.96, p-value=0.026). (**Table 3**)

**Table 3:**
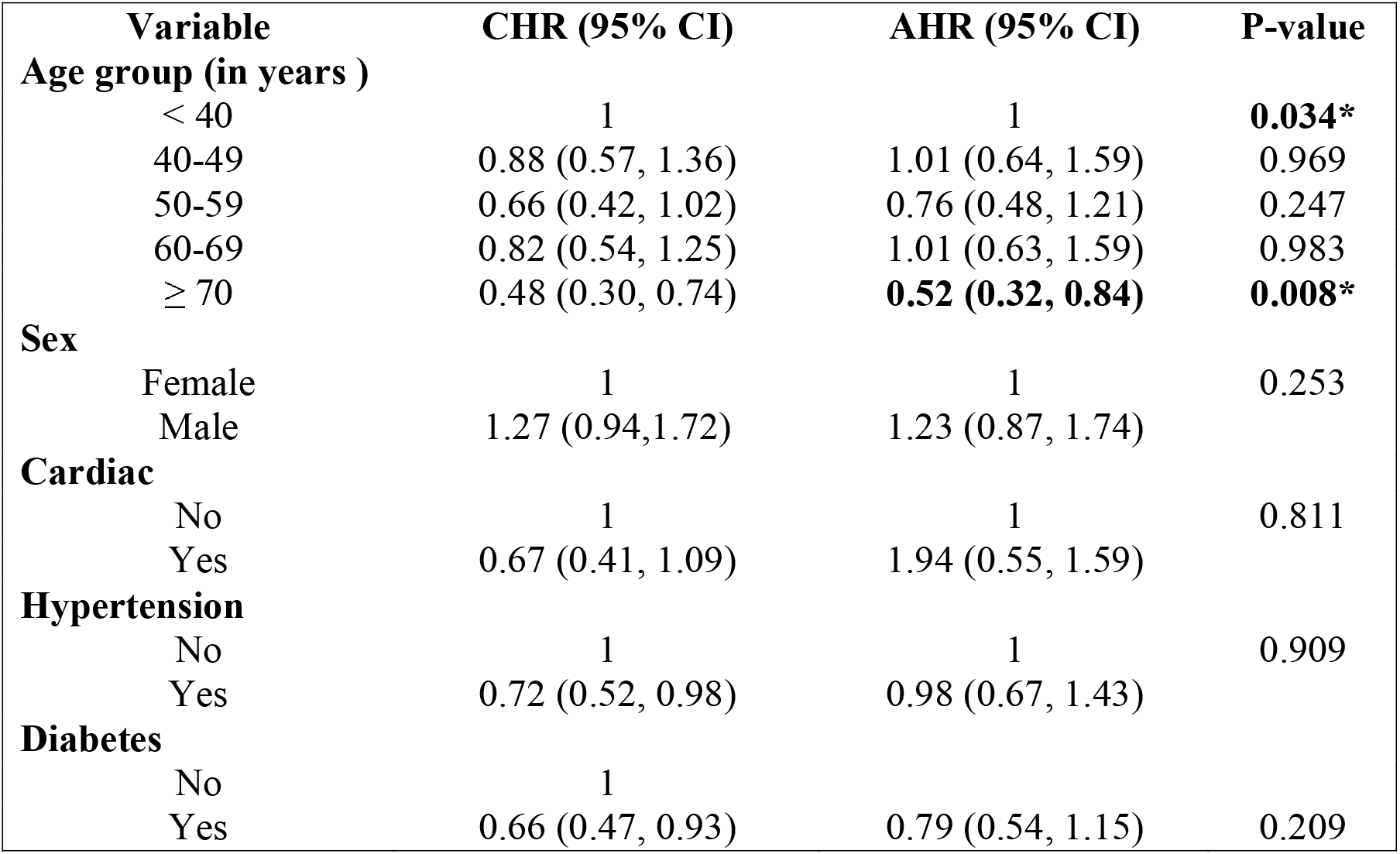
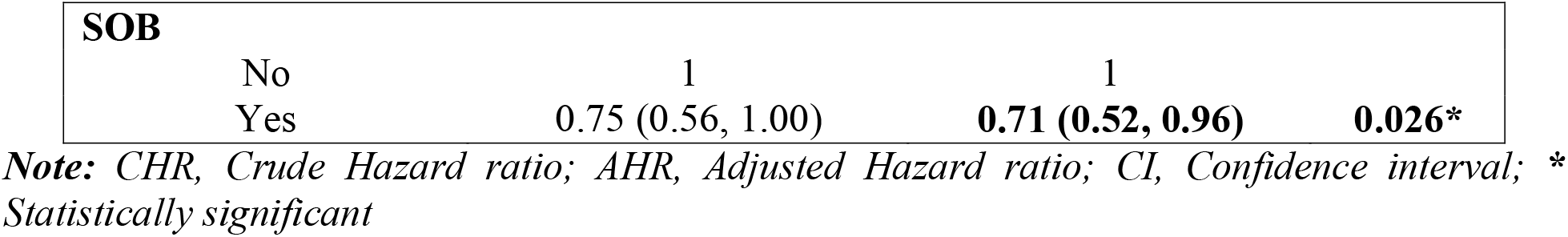
Result of Multivariable Cox proportional hazard model among COVID-19 patients (n=244)

| Variable | CHR (95% CI) | AHR (95% CI) | P-value |
| --- | --- | --- | --- |
| **Age group (in years )** |  |  |  |
| < 40 | 1 | 1 | **0.034\*** |
| 40-49 | 0.88 (0.57, 1.36) | 1.01 (0.64, 1.59) | 0.969 |
| 50-59 | 0.66 (0.42, 1.02) | 0.76 (0.48, 1.21) | 0.247 |
| 60-69 | 0.82 (0.54, 1.25) | 1.01 (0.63, 1.59) | 0.983 |
| $\geq 70$ | 0.48 (0.30, 0.74) | **0.52 (0.32, 0.84)** | **0.008\*** |
| **Sex** |  |  |  |
| Female | 1 | 1 | 0.253 |
| Male | 1.27 (0.94, 1.72) | 1.23 (0.87, 1.74) |  |
| **Cardiac** |  |  |  |
| No | 1 | 1 | 0.811 |
| Yes | 0.67 (0.41, 1.09) | 1.94 (0.55, 1.59) |  |
| **Hypertension** |  |  |  |
| No | 1 | 1 | 0.909 |
| Yes | 0.72 (0.52, 0.98) | 0.98 (0.67, 1.43) |  |
| **Diabetes** |  |  |  |
| No | 1 |  |  |
| Yes | 0.66 (0.47, 0.93) | 0.79 (0.54, 1.15) | 0.209 |
| **SOB** |  |  |  |
| No | 1 | 1 |  |
| Yes | 0.75 (0.56, 1.00) | **0.71 (0.52, 0.96)** | **0.026\*** |

*Note: CHR, Crude Hazard ratio; AHR, Adjusted Hazard ratio; CI, Confidence interval; \* Statistically significant*

## DISCUSSION

The study aimed at assessing the duration of supplemental oxygen requirement and identifying its predictors. Assessing this outcome as an indicator of clinical improvement from the disease is important as measuring the duration of oxygen requirement assists in deciding on building hospitals capacity in terms of oxygen facility arrangement and organizing ICU and wards with inbuilt supplemental oxygen system so that maximum capacity can be used to provide better care to patients in the country. Accordingly, the median time to getting off supplemental oxygen therapy among the studied population was 6 days (IQR, 4.3-20.0). On the univariate analysis, age group, sex, cardiac disease, hypertension, diabetes mellitus and shortness of breath were found to be independent predictors of duration of oxygen requirement among COVID-19 patients. But on the multivariable Cox proportional hazard model, after adjusting for other covariates, only age group and shortness of breath were found to be significant predictors of duration of supplemental oxygen requirement among COVID-19 patients.

Accordingly, after adjusting for other covariates, the rate of getting off supplemental oxygen therapy among patients 70 years and older was 47.8% lower than patients less than 40 years old. This implies that the time needed to get off supplemental oxygen therapy was significantly longer among older patients compared with the younger patients. This could be because of the increased risk of having concomitant comorbid illness and the normal aging process that diminishes body’s defense mechanism. These factors result in a more severe disease progression with delayed recovery or death. Studies also support this finding showing that old age is associated with high risk of developing symptomatic disease, severe disease category and death from COVID-19 as compared to younger age group. Especially the patients 70 years and older were found to be vulnerable to much worse disease progression and outcome than other age groups (19-22).

Having a subjective complaint of shortness of breath at admission was found to be associated with a significantly prolonged duration of supplemental oxygen requirement. The rate of getting off supplemental oxygen therapy among patients with shortness of breath was 29.5% lower compared to those patients with no such complaint on admission. Shortness of breath is a sign of lung disease and SARS-COV-2 can affect any part of the body system but the lungs are said to be more susceptible because the virus entry in to the body is made through the airways. The other reason could be the abundance of angiotensin-converting enzyme 2 in the lungs that is used as a receptor by the SARS-COV-2 to enter in to the body cells. Also, if the individual develops pneumonia, it is usually going to affect both lungs compromising the lungs capacity and resulting in a drop in blood oxygen level. Because of these reasons, if the lungs got hit by the virus, it results in a more severe disease especially among those with underlying pulmonary disease resulting in a severe disease presentation and delayed recovery causing a prolonged oxygen requirement.

On the other hand, the result shows that the duration of oxygen requirement doesn’t significantly differ based on sex and co-morbid illness/s history. In conclusion, the average duration of supplemental oxygen therapy requirement among COVID-19 patients was 6 days. This can be used as a guide in planning institutional oxygen requirement, bed demand at Intensive care unit and wards with inbuilt oxygen supply system, and in predicting patient turn over at these units. This in turn can be used to predict institutional capacity to admit and treat patients who require oxygen therapy.

Age group of 70 years and older and having a subjective complaint of shortness of breath were found to be associated with prolonged duration of supplemental oxygen therapy requirement. This implies that, earlier identification of disease progression is advised to identify these groups of patients so that early intervention and maximum care can be provided to prevent complication from the disease and the supplemental oxygen therapy itself.

In this study the role of laboratory and radiologic parameters were not assessed as data was not fully available for all patients. We recommend further study including these parameters in addition to the clinical criteria so that the role of these parameters on the disease can be understand better and can be utilized as disease course predicting variables.

## Data Availability

All relevant data are available upon reasonable request.

## Acknowledgment

The authors would like to thank St. Paul’s Hospital Millennium Medical College for facilitating the research work.

## Declaration

### Authors Contribution

TWL and ISH conceived the study. TWL designed the study, revised data extraction sheet, performed statistical analysis, and drafted the initial manuscript. All authors contributed to the conception of the study and obtained patient data. All authors undertook review and interpretation of the data. All authors revised the manuscript and approved the final version.

### Availability of data and materials

All relevant data are available upon reasonable request.

## REFERENCES

1. World Health Organization. WHO Director-General’s opening remarks at the media briefing on COVID19. March, 2020.

2. Health EFMo. National COVID-19 Daily report. January 25, 2020.

3. Ethiopian Federal Ministry of Health. Covid19 Management Handbook. 2020.

4. World Health Organization. Clinical management of severe acute respiratory infection (SARI) when COVID-19 disease is suspected: Interim guidance.. Geneva: March 2020

5. Bhatraju PK, Ghassemieh BJ, Nichols M, et al. Covid-19 in critically ill patients in the Seattle region?case series. N Engl J Med. 2020;382(21):2012–22.

6. Cai Q, Huang D, Ou P, et al. COVID-19 in a designated infectious diseases hospital outside Hubei Province, China. Allergy. 2020.

7. Cao B, Wang Y, Wen D, et al. A trial of lopinavir-ritonavir in adults hospitalized with severe covid-19. N Engl J Med. 2020;382:1787–799.

8. Guan Wj, Ni Zy, Hu Y, et al. Clinical characteristics of coronavirus disease 2019 in China. N Engl J Med. 2020.

9. Inciardi RM, Adamo M, Lupi L, et al. Characteristics and outcomes of patients hospitalized for COVID-19 and cardiac disease in Northern Italy. Eur Heart J. 2020;41(19):1821–9.

10. Intensive Care National Audit and research Centre (ICNArC). Report on 2249 patients critically ill with COVID-19. 2020.

11. Richardson S, Hirsch JS, Narasimhan M, et al. Presenting characteristics, comorbidities, and outcomes among 5700 patients hospitalized with COVID-19 in the New York City area. JAMA Intern Med. 2020;323(20):2052–9.

12. Shi H, Han X, Jiang N, et al. Radiological findings from 81 patients with COVID-19 pneumonia in Wuhan, China: a descriptive study. Lancet Infect Dis. 2020;20(4):425–34.

13. Kaijin Xu, Yanfei C, Jing Y, et al. Factors Associated With Prolonged Viral RNA Shedding in Patients with Coronavirus Disease 2019 (COVID-19) Clinical infectious diseases. 2020;2020.

14. Lin Qi YY, Dixuan J,et al. Factors associated with the duration of viral shedding in adults with COVID-19 outside of Wuhan, China: a retrospective cohort study. International journal of infectious diseases. 2020;96 (2020):531–7.

15. Xiaowen Hu, Yuhan X, Jing J, et al. Factors associated with negative conversion of viral RNA in patients hospitalized with COVID-19 Science of the Total Environment,. 2020;728 (2020).

16. Yingying Lu, Yi Li, Wenyue D, et al. Symptomatic Infection is Associated with Prolonged Duration of Viral Shedding in Mild Coronavirus Disease 2019: A Retrospective Study of 110 Children in Wuhan. Pediatr Infect Dis J. July 2020;39(7):95–9.

17. Zhou B, She J, Wang Y, Ma X. The duration of viral shedding of discharged patients with severe COVID-19. Clin I n f e c t Dis 2020 Apr 17.

18. David Hosmer and Lemeshow. Applied survival analysis. 2nd edition ed 2008.

19. Du R-H, Liang L-R, Yang C-Q, et al. Predictors of mortality for patients with COVID-19 pneumonia caused by SARSCoV-2: a prospective cohort study. Eur Respir J. 2020;2020(55: 2000524).

20. Gupta S, Hayek SS, Wang W, et al. Factors Associated With Death in Critically Ill Patients With Coronavirus Disease 2019 in the US. JAMA Intern Med.2020;e203596.

21. Williamson E.J., Walker A.J., Bhaskaran K, et al. Factors associated with COVID-19-related death using OpenSAFELY. Nature.584:430–6 (2020).

22. Xiaochen Li, Shuyun Xu, Muqing Yu, et al. Risk factors for severity and mortality in dult COVID-19 inpatients in Wuhan. J ALLERGY CLIN IMMUNOL. 2020;146:110–8.

